# Polygenic scores for tobacco use provide insights into systemic health risks in a diverse EHR-linked biobank in Los Angeles

**DOI:** 10.1101/2023.02.05.23285493

**Authors:** Vidhya Venkateswaran, Kristin Boulier, Yi Ding, Ruth Johnson, Arjun Bhattacharya, Bogdan Pasaniuc

## Abstract

**Importance:** Tobacco use is heavily influenced by environmental factors with significant genetic contributions. An extensive evaluation of the genetic variants predisposing to tobacco use is necessary to understand associated health risks and formulate equitable health policies.

**Objective:** To evaluate the predictive performance, risk stratification, and potential systemic health effects of tobacco use disorder (TUD) predisposing germline variants using a European-derived polygenic score (PGS) in the UCLA ATLAS biobank.

**Design, Setting, and Participants:** Publicly available TUD-PGS, developed in European ancestry individuals, was evaluated in participants enrolled in the UCLA ATLAS biobank - a multi-ancestry, hospital-based biobank with participant genotypes linked to their de-identified medical records.

**Main Outcomes and Measures:** The outcomes of interest were (a) tobacco use disorder and (b) 1,847 phenotypes, identified by phecodes (aggregated ICD codes) extracted from electronic health records.

**Results:** Among genetically inferred ancestry groups (GIAs), TUD-PGS associated with TUD in European American (EA) (OR: 1.20, CI: [1.16, 1.24]), Hispanic/Latin American (HL) (OR:1.19, CI: [1.11, 1.28]), and East Asian American (EAA) (OR: 1.18, CI: [1.06, 1.31]) GIAs but not in African American (AA) GIA (OR: 1.04, CI: [0.93, 1.17]). Similarly, TUD-PGS offered strong risk stratification across quantiles in the EA and HL GIAs but inconsistently in EAA and AA GIAs. In a cross-ancestry phenome-wide association meta-analysis, TUD-PGS was associated with cardiac, respiratory, psychiatric, and metabolic phecodes (17 phecodes at P < 2.57e-5). In individuals with no history of smoking (never-smokers), the top TUD-PGS associations were obesity and alcohol-related disorders (P = 3.54E-07, 1.61E-06). Mendelian Randomization (MR) analysis provides evidence of a causal association between tobacco use and adiposity measures.

**Conclusions and Relevance:** This study provides an investigation of TUD-PGS across multiple ancestries and a range of phenotypes in a hospital-based biobank, suggesting shared biological pathways between tobacco use, alcohol use disorder, and obesity. European ancestry TUD-PGS demonstrated inconsistent performance in non-European ancestries for risk prediction and stratification. Equitable clinical translation of TUD-PGS will require the inclusion of multiple ancestry populations at all levels of TUD genetic research. Additionally, TUD-predisposed individuals may require comprehensive tobacco use management approaches to address underlying addictive tendencies.

**Key Points:** *Question:* How does a European-derived polygenic score (PGS) for tobacco use disorder (TUD) perform in ancestrally diverse individuals within a real-world medical system and what are the systemic health risks in TUD predisposed individuals?

*Findings:* European ancestry TUD-PGS performs inconsistently in non-European ancestry populations. TUD-PGS correlates with cardiometabolic, respiratory, and psychiatric phenotypes. In individuals with no history of smoking, TUD-PGS correlates with obesity and alcohol-related disorders.

*Meaning:* Existing TUD-PGS is not generalizable across ancestries. TUD-predisposed individuals are at risk for obesity and alcohol use disorder in the absence of smoking behavior, suggesting shared genetic etiology between these phenotypes.

## Introduction

Tobacco use is a leading cause of global mortality and morbidity, contributing to several systemic conditions, including cardiometabolic diseases and cancers^1,2^. Tobacco use is heavily influenced by environmental factors^3^, with growing evidence of underlying contributions from genetic factors^4,5^. Multi-ancestry studies on tobacco use behaviors report an estimated heritability (i.e., the proportion of the phenotypic variance explained by genetics) of smoking behaviors ranging between 5-18%^4,5^. Tobacco use management can greatly benefit from precision medicine initiatives, with the inclusion of baseline genetic risk to develop individualized preventive and therapeutic strategies for tobacco use cessation. These efforts require a thorough understanding of the effects of a genetic predisposition to tobacco use across ancestry groups and the impact of tobacco predisposition on the overall systemic health of an individual.

Genome-wide association studies (GWAS) are a commonly used approach to identify genetic variants associated with complex traits. GWAS have identified >2000 loci associated with tobacco use traits such as smoking behaviors and nicotine dependence^4,5^. Polygenic scores (PGS) are composite scores that capture the overall genetic predisposition to a trait (e.g., tobacco use) by aggregating the weighted effects for multiple variants of interest, thus capturing a larger proportion of phenotypic variation. Polygenic scores have been used in research for disease prediction and to evaluate disease correlations^6^. Tobacco PGS, in particular, have been associated with diseases such as schizophrenia, and substance use disorders^7–11^.

Phenome-wide association studies (PheWAS) systematically test the association of a single genetic variant across multiple phenotypes^12^. These phenotypes are identified using ‘Phecodes’ which are ICD-codes that are aggregated to be clinically meaningful. Thus, a PheWAS can identify variant-phecode associations across a spectrum of phenotypes. Tobacco use polygenic scores can be combined with a PheWAS to create a PGS-PheWAS, a powerful way to examine the potential pleiotropic effects of multiple genetic variants that predispose to tobacco use disorder and identify systemic disease risks for individuals with a genetic predisposition to tobacco use^13^.

In this study, we used a publicly available PGS for tobacco use disorder, developed in European-ancestry individuals in UK Biobank^14,^ and imputed these scores into the UCLA ATLAS biobank ^15–19^. We found that the TUD-PGS demonstrated inconsistent predictive performance and risk stratification in non-European ancestry groups. In a PGS-PheWAS, we detected several phecodes associated with a genetic predisposition to tobacco use, mainly cardiometabolic, respiratory, and neuropsychiatric phenotypes. Stratifying to individuals with no history of smoking, we identified TUD-PGS associations with obesity and alcohol-related disorders. Finally, we used publicly available GWAS summary statistics to perform Mendelian randomization^20^ analysis and found evidence of causality between tobacco use and adiposity measures.

## Methods

### Study population

All analyses were performed with UCLA ATLAS Biobank data - a biobank embedded within the UCLA Health medical system^15–19^. UCLA Health is a comprehensive healthcare system serving the population in and around the greater Los Angeles area. The UCLA Institute for Precision Health is home to the UCLA ATLAS biobank with >40k participants genotyped, of which 26,517 participants were included in this study. This large-scale collection of genotyped biospecimens is integrated with the UCLA Data Discovery Repository (DDR), containing de-identified patient electronic health records (EHR) which include clinical, procedural, laboratory, prescription, and demographic information.

Final analyses included all ATLAS participants with complete information on the outcome and covariates (described below in detail). For ancestry-specific analysis, we included European American, Hispanic/Latin American, East Asian American, and African American ancestry groups with sufficient sample sizes for analysis (n = 17752, 4599, 2603, and 1563).

### Data Processing and Population Stratification

Detailed information on data processing can be found in previous publications^15–19^ Blood samples were collected from consented participants and genotyped on a custom array^21^. Initial array-level quality control measures included removing strand ambiguous SNPs and variants with >5% missingness. After restricting to unrelated individuals, the QC-ed genotypes were imputed to the TOPMed Freeze5 reference using the Michigan Imputation Server^22,23^. The final QC steps were to filter the variants at the threshold of R^2^ > 0.90 and minor allele frequency > 1%. All quality control steps were conducted using PLINK 1.9^24^.

We computed the top 10 principal components for our study population using FlashPCA2 software^25^ and grouped our study population into genetically inferred ancestry groups (GIAs) - European American (EA), Hispanic/Latin American (HL), East Asian American (EAA), African American (AA) by k-nearest neighbor (KNN) stratification of the principal components, using the continental ancestry populations from 1000 Genomes Project^26,27^. To account for differences in population stratification across GIA groups, for the PGS-PheWAS analysis, we conducted individual PGS-PheWAS within each GIA group and then meta-analyzed across GIA groups to obtain cross-ancestry results.

### Polygenic Score Imputation within ATLAS

We used a publicly available polygenic score trained on 391,124 European individuals from the UK biobank for the trait ‘tobacco use disorder’ from the PGS catalog (PGS002037)^14,28^. This PGS was selected for two reasons: (1) the PGS was trained on the same phecode for TUD that is available in ATLAS and (2) there is a high degree of overlap with ATLAS genotyped variants (802,624 of 847,691 total variants in TUD-PGS overlapping with ATLAS data). The original PGS training analyses were performed using LDpred2^29^ and adjusted for the following covariates: sex, age, birth date, Townsend’s deprivation index, and the first 16 principal components of the genotype matrix. We computed the PGS for each ATLAS participant by multiplying the individual risk allele dosages by their corresponding weights that are provided by the PGS catalog^28^. The PGS was mean-centered and standardized by the standard deviation within each GIA group to generate a PGS Z-score.

### Phecodes

ICD9 and ICD10 billing codes were aggregated into clinically meaningful groupings called phecodes. These phecode groupings used mappings derived from the PheWAS catalog, v1.2^30^. Cases were defined by the presence of an ICD code tagged by the respective phecode and controls by the absence of the ICD codes. Tobacco use disorder diagnosis was derived from the presence of “tobacco use disorder” phecode (318.00) within an individual’s health record which groups ICD codes (F17.200, F17.201, F17.210, F17.211, F17.220, F17.221, F17.290, F17.291, O99.33, O99.330, O99.331, O99.332, O99.333, O99.334, O99.335, Z87.891) for tobacco use disorder (TUD). For the PheWAS analysis, we used 1847 phecodes, extracted from each individual’s health record as described above, to capture phenotypes across the phenotypic spectrum^30^.

### Statistical Analysis

All analysis was conducted in either Python 2.6.8^31^ or R 4.2.1^32^.

#### a) Predictive performance and Risk Stratification

We analyzed the predictive performance of the standardized TUD-PGS across ancestry groups and risk quantiles using GIA-stratified logistic regression models, with the phecode for TUD as the outcome and with predictors including terms for age, sex, the first five principal components of the genotype matrix, and insurance class.

We include insurance class information as the closest proxy to bias introduced by participation and access to healthcare within the de-identified electronic health records^33^. This insurance class variable consists of the type of insurance used by the patient in their clinical encounters. The classes include - “Public”, “Private” or “Self-pay”. Public class includes ‘Medicare’, ‘Medicare Advantage’, ‘Medicare Assigned’, ‘Medi-Cal’, ‘Medicaid’, and ‘Medi-Cal Assigned’.

Private class includes - ‘International Payor’, ‘Group Health Plan’, ‘Worker’s Comp’, ‘Tricare’, ‘UCLA Managed Care’, ‘Blue Shield’, ‘Commercial’, ‘Blue Cross’, ‘Package Billing’ and ‘Other’.

Odds ratios were calculated within each GIA, with P-values from Wald-type test statistics and a Bonferroni-corrected significance level of 0.0125 = (0.05/4). For risk stratification analysis, we grouped individuals of each GIA group into 5 groups of equal size based on their PGS and compared the quintile with the highest score with the quintile with the lowest scores.

#### b) Phenome-wide association meta-analysis

For the phenome-wide association analysis, we tested the association between the standardized TUD-PGS and 1847 electronic health record-derived phecodes across the phenome. Each GIA-specific PheWAS analysis consisted of logistic regressions across 1847 EHR-derived phecodes, controlling for age, sex, first 5 PCs, and insurance class. For the cross-ancestry meta-analysis, we use the PGS-PheWAS results computed within each GIA group and meta-analyze across these ancestry groups using a random effect, inverse variance weighted model using the metafor (version 3.4) package in R^34^. We use a phenome-wide Bonferroni-corrected p-value significance threshold of 2.7e-05 to adjust for the multiple testing burden (P = 0.05/1847 tests for each trait identified by phecodes). The never-smoker analysis followed a similar analysis plan, restricted to individuals of European American GIA with no history of smoking recorded by their provider within their medical records (n=9,921).

#### c) Mendelian Randomization

We evaluated causality using Mendelian Randomization (MR) methods to test for and evaluate the causality between tobacco use and obesity^20^. We used summary statistics from GSCAN Consortium GWAS for “Cigarettes Smoked Per Day” (249,752 males and females of European Ancestry and 12,003,613 SNPs)^35^ and summary statistics from MRC Integrative Epidemiology Unit - the University of Bristol and UKBB GWAS for “Waist Circumference” (462,166 males and females of European Ancestry and 9,851,867 SNPs)^36^ as our instrumental variables to test the causal association between tobacco use and obesity measures. We performed a second MR analysis to validate the previous analysis using summary statistics for ‘Body Mass Index - BMI’ using summary statistics from UK Biobank^36^ (461,460 individuals and 9,851,867 SNPs), using the same ‘Cigarettes smoked per day’ summary statistics from GSCAN as the outcome. We used the ‘TwoSampleMR’ R package to extract instruments, harmonize and obtain effect sizes from multiple MR methods (MR Egger, Weighted median, Inverse variance weighted, Simple mode, and Weighted mode)^37^.

## Results

### Baseline characteristics of cases and controls in ATLAS Biobank

The final analysis included n = 26,517 individuals with complete information on all covariates. Within the “TUD” phecode, our study population consisted of 8,667 cases and 17,850 controls. The average age of individuals with a TUD phecode was 64.2 years. Participant sex was significantly associated with TUD phecode with 56% of the phecode represented by the male sex. Four genetically inferred ancestry groups had sufficient sample size to perform the analyses - European American (EA), Hispanic/Latin American (HL), East Asian American (EAS), and African American ancestry (AA) (n=17,752, 4,599, 2,603, and 1,563). (***Table 1***).

**Table 1.** Baseline characteristics of ATLAS participants included in this study.

|  |  | **Overall** |
| --- | --- | --- |
| **n** |  | 26517 |
| **Patient Age, median [Q1, Q3]** |  | 62.0 [47.0,72.0] |
| **Sex, n (%)** | Female | 14425 (54.4) |
|  | Male | 12092 (45.6) |
| **Insurance Class, n (%)** | Private | 16207 (61.1) |
|  | Public | 9469 (35.7) |
|  | Self-Pay | 841 (3.2) |
| **Tobacco Use Disorder, n (%)** | Controls | 17850 (67.3) |
|  | Cases | 8667 (32.7) |
| **Genetically Inferred Ancestry, n (%)** | African American (AA) | 1563 (5.9) |
|  | Hispanic/Latin American (HL) | 4599 (17.3) |
|  | East Asian American (EAA) | 2603 (9.8) |
|  | European American (EA) | 17752 (66.9) |

### TUD-PGS predicted and risk-stratified strongly for TUD in EA, HL GIAs, and inconsistently in EAA and AA GIAs across risk quantiles

The TUD-PGS associated with the phecode for TUD within the ATLAS biobank for individuals of European American (EA) GIA (OR:1.19, CI: [1.11, 1.28]), Hispanic/Latin American (HL) GIA (OR:1.21, CI: [1.12, 1.31]), and East Asian American (EAA) GIA groups (OR: 1.18, CI: [1.06, 1.31]). TUD-PGS was not associated with TUD in individuals of African American (AA) GIA group (OR: 1.04, CI: [0.93, 1.17]). ***(Supp Table 1)***

To evaluate the risk stratification provided by TUD-PGS for tobacco use disorder, we evaluated PGS effect sizes across PGS quintiles. When compared to the quintile with the lowest score, the quintile with the highest polygenic scores demonstrated an OR = 1.69 (CI: [1.51, 1.88]) in EA, and 1.71 (CI: [1.36, 2.14]) in HL. The TUD-PGS offered strong risk stratification for individuals of EA GIA and for the top two risk quintiles in HL. Risk stratification was weaker and inconsistent in the EAA, (OR = 1.60, CI = [1.15, 2.24]) and AA ancestry groups (OR = 1.02, CI = [0.71, 1.47]). (***Fig 1, Supp Table 2***)

**Figure 1.**
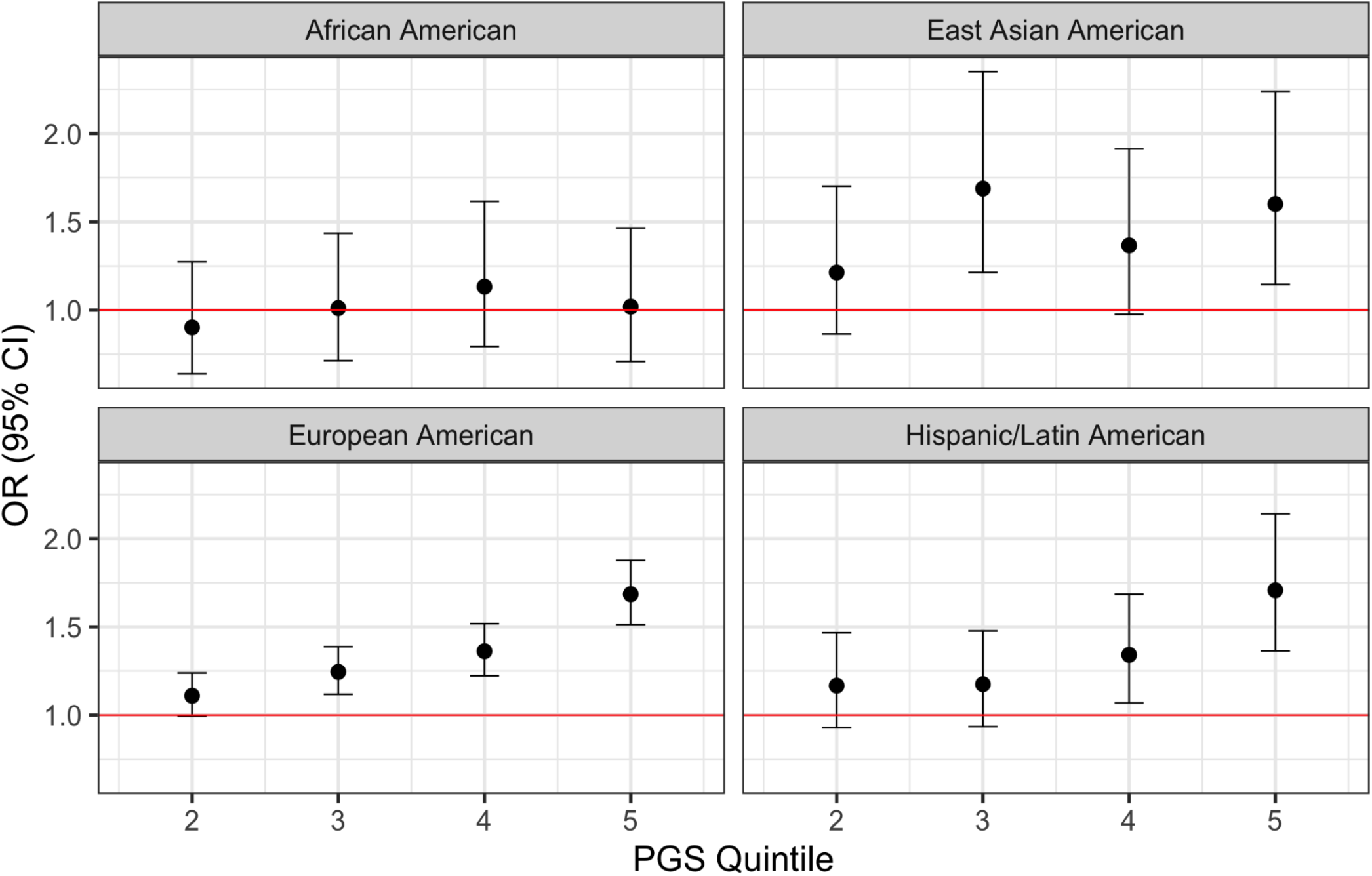
TUD-PGS correlates with TUD phecode in EA, HL, and EAA ancestries across risk quintiles. The X-axis represents the top 4 quintiles grouped according to TUD-PGS. Y axis represents effect sizes represented by odds ratios. The red line indicates OR = 1. Effect sizes between TUD-PGS and TUD phecode vary across PGS-quintiles in 4 genetically inferred ancestry groups with strong risk stratification noted in EA and HL and inconsistent risk stratification in AA and EAA groups.

### TUD-PGS replicated clinically observed phenotypic associations with TUD in PGS-PheWAS

In a PheWAS of the TUD-PGS across 1847 phecodes (***Supp Fig 1a***), meta-analyzed across 4 GIAs, we found 17 significant associations at Bonferroni-adjusted P < 0.05 after adjusting for age, sex, first 5 principal components, and health insurance information. The top phecodes associated with the TUD-PGS were ‘morbid obesity’, ‘obstructive chronic bronchitis’, ‘substance addiction and disorders’, and ‘ischemic heart disease’ (P = 1.38E-09, 2.73E-09, 4.45E-08, 1.61E-07) (***Fig 2a***). Phecode groups with multiple associations were circulatory (n=5), respiratory (n=3), neurological (n=2), and metabolic (n=2) phenotypes (***Supp Table 3)***

**Figure 2a.**
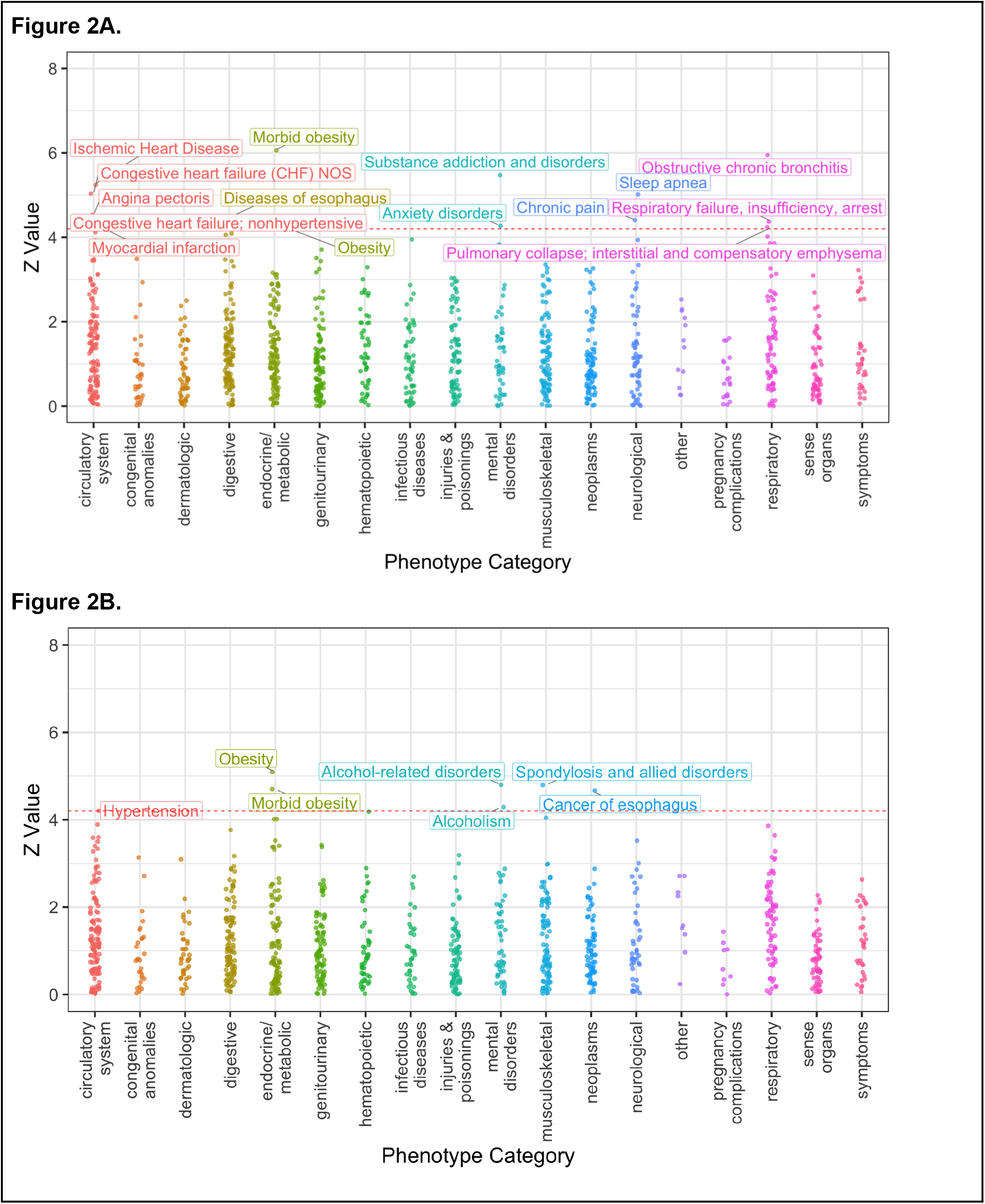
TUD-PGS-PheWAS plot across 1847 phecodes (cross-ancestry meta-analysis) Associations between TUD-PGS and 1847 phecodes across the phenome, meta-analyzed across 4 GIA groups with significant associations labeled. The X-axis represents the Z value (beta/SE). Each color represents a phecode category and each dot represents a phecode. Phenome-wide significance is represented by the red dashed line at a Z value = 4.2 which corresponds to a P value of 2.57e-5 (1847 tests/0.05). Top associations were noted in circulatory, metabolic, mental and respiratory phenotype categories. **Figure 2b TUD-PGS-PheWAS plot across 1847 phecodes in never-smokers of EA ancestry group** Associations between TUD-PGS and 1847 phecodes across the phenome in never-smokers of EA ancestry with significant associations labeled. The X-axis represents the Z value (beta/SE). Each color represents a phecode category and each dot represents a phecode. Phenome-wide significance is represented by the red dashed line at a Z value = 4.2 which corresponds to a P value of 2.57e-5 (1847 tests/0.05). In TUD-PGS-PheWAS restricted to ‘never smokers’, top associations were obesity and alcohol-related disorders.

### TUD-PGS associated with alcohol use and obesity phecodes in the absence of smoking behavior in PGS-PheWAS

We repeated the PGS-PheWAS association analysis, restricting to “never-smokers” in individuals of EA ancestry - i.e. individuals who reported to their provider that they have never smoked tobacco (***Supp Fig 1b***). In this analysis, the TUD-PGS demonstrated associations with obesity, alcohol-related disorders, cancer of the esophagus, and hypertension (P = 3.54E-07, 1.61E-06, 3.05E-06, 2.62E-05) (***Figure 2b, Supp Table 4***).

In an evaluation of the trends of obesity and alcohol-related disorders across quintiles of the TUD-PGS, we observed higher ORs among never-smokers when compared to ever-smokers for obesity and alcohol-related disorders. TUD-PGS offered inconsistent risk stratification for obesity and alcohol-related disorders in ever-smokers (***Figure 3***). In contrast, a reverse trend is noted in lung cancer, an established trait associated with smoking behavior, where we observed higher ORs in ever-smokers compared to never-smokers. (***Supp figure 2***)

**Figure 3.**
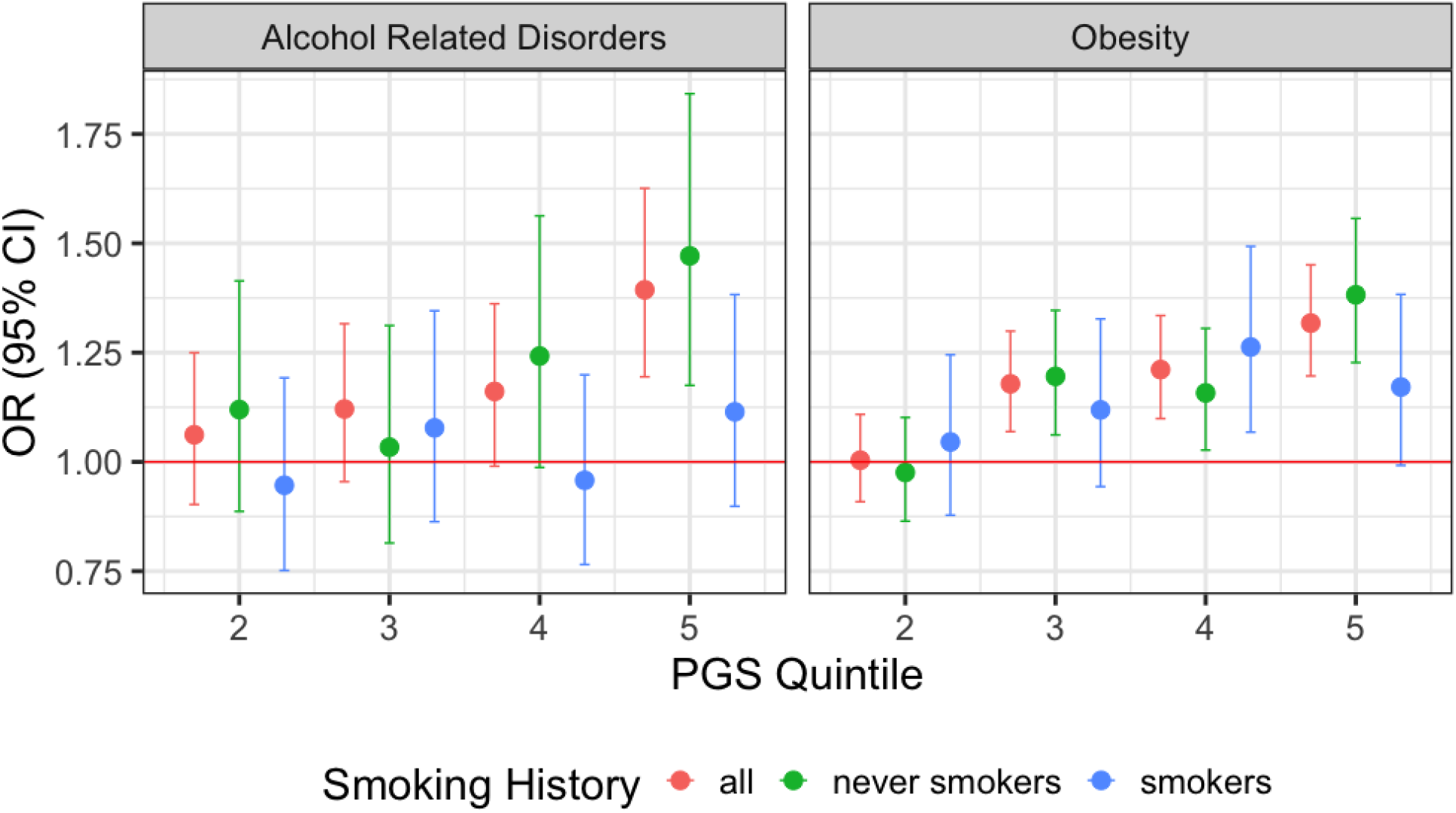
TUD-PGS associations with Alcohol-related disorders and Obesity among all vs ever vs never-smokers across TUD-PGS quintiles. Associations between TUD-PGS quintiles and Alcohol-related disorders (phecode = 317.0) and Obesity (phecode = 278.1). The X-axis represents the top 4 quintiles grouped according to TUD-PGS. Y axis represents effect sizes represented by odds ratios. The red line indicates OR =1. TUD-PGS risk-stratifies for the phecodes for alcohol-related disorders and obesity in ‘never smokers’ but not in ‘ever smokers’.

### Mendelian randomization analysis finds evidence of causality in the association between obesity and tobacco use

To evaluate if the association between obesity and tobacco use is causal, we performed Mendelian randomization (MR) analysis between two quantitative measures of obesity and tobacco use using the phenotypes “waist circumference” and “cigarettes smoked per day”. From the results of multiple MR methods, we observed that the exposure - ‘waist circumference’ demonstrated significant positive causal associations with the outcome - ‘cigarettes smoked per day’ across all methods used to test this association. (MR Egger, Weighted median, Inverse variance weighted, Simple mode, Weighted mode with P = 2.39E-03, 1.50E-32, 1.49E-46, 8.22E-05, 2.05E-08 respectively). A second MR analysis of ‘body mass index’ and ‘cigarettes smoked per day’ provided validation for this causal association - (MR Egger, Weighted median, Inverse variance weighted, Simple mode, Weighted mode P = 2.65E-03, 8.34E-33, 1.17E-45, 8.23E-06, 5.78E-07). ***Supp Figure 3*** presents causal effect estimates and confidence intervals.

## Discussion

In this study, we examined the predictive performance and risk stratification of a publicly available, European ancestry PGS for tobacco use disorder in a multi-ancestry EHR-linked biobank. Our results demonstrated that this TUD-PGS predicts TUD and risk-stratified European American GIA and Hispanic/Latin GIA groups. However, inconsistent prediction and risk stratification was noted in the East Asian American and African American GIA groups.

Based on these results, we anticipate two issues if TUD-PGS is used clinically to identify individuals at high risk for tobacco use. First, the risk stratification offered only to certain ancestry populations does not allow for equitable translation of genetic research. Second, the application of these PGS to individual-level clinical decisions must proceed with caution with additional extensive validation with clinical history. At present, we do not recommend interventions solely based on being classified as “high risk” by TUD-PGS due to large uncertainty in imputed polygenic scores at an individual level^38^ and inconsistent performance in non-European populations.

Next, we evaluated the potential pleiotropic effects of TUD predisposing variants using the PGS to conduct a phenome-wide association analysis. Additionally, we repeated this analysis in a subgroup of participants without a reported history of smoking behavior, to evaluate the systemic associations of a genetic predisposition to tobacco use in the absence of tobacco use behavior. The PGS-PheWAS meta-analysis demonstrated significant associations with respiratory and cardiovascular phenotypes, both of which have robust clinical and biological evidence^39,40^. Other significant associations were in the category of psychiatric disorders, including associations with anxiety disorders and substance addiction disorders. These psychiatric disorder associations have been consistently reported in past genetic studies of smoking and tobacco use^41^.

In the analysis of ‘never-smokers’, phenotypes associated with tobacco use behaviors, namely, respiratory and cardiovascular phecodes, did not demonstrate statistical significance. Instead, we observed associations with psychiatric phecodes including alcohol-related disorders, and metabolic phecodes such as obesity. The MR analysis results suggest a causal association between tobacco use and adiposity, in line with other published literature with similar directionality and effect sizes^42^. Together, the associations between tobacco use, obesity, and alcoholism are suggestive of shared genetic architecture between these traits, likely originating from the biological regulation of impulsivity and addictive behaviors^43^.

Clinically, these findings will have implications for patients with tobacco use disorder: in the presence of a genetic propensity for TUD, we demonstrate that the individual may be at risk for obesity and alcohol use disorder even if tobacco use behavior is absent. For patients in the high-risk genetic propensity to TUD group, these findings would shift the focus of the therapeutic strategy from tobacco cessation to a more comprehensive regulation of biological pathways that underlie addiction and impulsivity.

A major strength of this study is that we evaluated TUD-PGS in an information-rich biobank across multiple genetically inferred ancestry groups. The rich phenotypic information available in the biobank allowed us to test associations across the phenome in a hypothesis-free manner, allowing for discovery. Another strength of the paper is that we accounted for possible confounding bias introduced by participation/access to healthcare bias, which can arise from using data from a hospital-based biobank, by using an insurance class variable as a proxy marker for participation and access.

While our study included individuals from 4 GIAs, we suffer from lower sample sizes in non-European ancestries for certain analyses. With continued enrollment in the UCLA ATLAS biobank, we hope to increase our non-European sample sizes and evaluate differential genetic effects in these ancestries. Next, phecodes are derived from ICD codes which are billing codes and may not always capture the entire picture of the individual’s disease history. The interpretations of our analyses are within the limitations of these data. We note that the risk of having a phecode in the electronic health record does not accurately reflect the risk of having the disease. The former comes with biases, including access to healthcare. We have attempted to address this bias introduced by healthcare access by including an insurance class information variable. Nevertheless, this difference must be considered when applying these results to the general population.

## Conclusions

The results of our study have implications for public health and clinical approaches to the treatment of tobacco use disorder. A genetic propensity to tobacco use might also signal an increased risk of obesity and alcohol use disorders, necessitating a more comprehensive preventive and therapeutic strategy for such individuals. Given the growing evidence on health risks associated with obesity and tobacco use, our results suggest possible shared genetic etiology between these two risk factors, strengthening the argument that public health approaches must consider this shared risk while formulating interventions.

## Data Availability

All data produced in the present work are contained in the manuscript.

## Acknowledgments

We gratefully acknowledge the Institute for Precision Health, participating patients from the UCLA ATLAS Precision Health Biobank, UCLA David Geffen School of Medicine, UCLA Clinical and Translational Science Institute, and UCLA Health.

## References

1. World Health Organization. WHO Report on the Global Tobacco Epidemic, 2017. Geneva: World Health Organization, 2017.

2. National Center for Chronic Disease Prevention and Health Promotion (US) Office on Smoking and Health. The Health Consequences of Smoking—50 Years of Progress: A Report of the Surgeon General. Centers for Disease Control and Prevention (US); 2014. Accessed July 14, 2022. http://www.ncbi.nlm.nih.gov/books/NBK179276/

3. Caraballo RS, Rice KL, Neff LJ, Garrett BE. Social and Physical Environmental Characteristics Associated With Adult Current Cigarette Smoking. Prev Chronic Dis. 2019;16:180373. doi:10.5888/pcd16.180373

4. Evans LM, Jang S, Hancock DB, et al. Genetic architecture of four smoking behaviors using partitioned SNP heritability. Addict Abingdon Engl. 2021;116(9):2498–2508. doi:10.1111/add.15450

5. Genetic diversity fuels gene discovery for tobacco and alcohol use | Nature. Accessed December 15, 2022. https://www.nature.com/articles/s41586-022-05477-4#MOESM3

6. Lewis CM, Vassos E. Polygenic risk scores: from research tools to clinical instruments. Genome Med. 2020;12(1):44. doi:10.1186/s13073-020-00742-5

7. Ohi K, Nishizawa D, Muto Y, et al. Polygenic risk scores for late smoking initiation associated with the risk of schizophrenia. Npj Schizophr. 2020;6(1):1–7. doi:10.1038/s41537-020-00126-z

8. Al-Soufi L, Martorell L, Moltó MD, et al. A polygenic approach to the association between smoking and schizophrenia. Addict Biol. 2022;27(1):e13104. doi:10.1111/adb.13104

9. Deak JD, Clark DA, Liu M, et al. Alcohol and nicotine polygenic scores are associated with the development of alcohol and nicotine use problems from adolescence to young adulthood. Addiction. 2022;117(4):1117–1127. doi:10.1111/add.15697

10. Cooke ME, Clifford JS, Do EK, et al. Polygenic score for cigarette smoking is associated with ever electronic-cigarette use in a college-aged sample. Addiction. 2022;117(4):1071–1078. doi:10.1111/add.15716

11. Bray M, Chang Y, Baker TB, et al. The Promise of Polygenic Risk Prediction in Smoking Cessation: Evidence From Two Treatment Trials. Nicotine Tob Res Off J Soc Res Nicotine Tob. 2022;24(10):1573–1580. doi:10.1093/ntr/ntac043

12. Denny JC, Ritchie MD, Basford MA, et al. PheWAS: demonstrating the feasibility of a phenome-wide scan to discover gene-disease associations. Bioinformatics. 2010;26(9):1205–1210. doi:10.1093/bioinformatics/btq126

13. Pendergrass SA, Brown-Gentry K, Dudek S, et al. Phenome-Wide Association Study (PheWAS) for Detection of Pleiotropy within the Population Architecture using Genomics and Epidemiology (PAGE) Network. PLOS Genet. 2013;9(1):e1003087. doi:10.1371/journal.pgen.1003087

14. Privé F, Aschard H, Carmi S, et al. Portability of 245 polygenic scores when derived from the UK Biobank and applied to 9 ancestry groups from the same cohort [published correction appears in Am J Hum Genet. 2022 Feb 3;109(2):373]. Am J Hum Genet. 2022;109(1):12–23. doi:10.1016/j.ajhg.2021.11.008

15. Chang TS, Ding Y, Freund MK, et al. Pre-existing conditions in Hispanics/Latinxs that are COVID-19 risk factors. iScience. 2021;24(3). doi:10.1016/j.isci.2021.102188

16. Lajonchere C, Naeim A, Dry S, Wenger N, Elashoff D, Vangala S, Petruse A, Ariannejad M, Magyar C, Johansen L, Werre G, Kroloff M, Geschwind D, An Integrated, Scalable, Electronic Video Consent Process to Power Precision Health Research: Large, Population-Based, Cohort Implementation and Scalability Study. J Med Internet Res 2021;23(12):e31121; doi: 10.2196/31121 : https://www.jmir.org/2021/12/e31121

17. Johnson R, Ding Y, Venkateswaran V, et al. Leveraging genomic diversity for discovery in an electronic health record linked biobank: the UCLA ATLAS Community Health Initiative. Genome Med. 2022;14(1):1–23. doi:10.1186/s13073-022-01106-x

18. Ruth Johnson, Yi Ding, Arjun Bhattacharya, Sergey Knyazev, Alec Chiu, Clara Lajonchere, Daniel H. Geschwind, Bogdan Pasaniuc, The UCLA ATLAS Community Health Initiative: Promoting precision health research in a diverse biobank, Cell Genomics, Volume 3, Issue 1, 2023, 100243, ISSN 2666-979X, https://doi.org/10.1016/j.xgen.2022.100243.

19. Naeim A, Dry S, Elashoff D, et al. Electronic Video Consent to Power Precision Health Research: A Pilot Cohort Study [published correction appears in JMIR Form Res. 2021 Oct 21;5(10):e33891]. JMIR Form Res. 2021;5(9):e29123. Published 2021 Sep 8. doi:10.2196/29123

20. Sanderson E, Glymour MM, Holmes MV, et al. Mendelian randomization. Nat Rev Methods Primer. 2022;2(1):1–21. doi:10.1038/s43586-021-00092-5

21. Infinium Global Screening Array-24 Kit | Population-scale genetics. Accessed January 31, 2023. https://www.illumina.com/products/by-type/microarray-kits/infinium-global-screening.html

22. Taliun D, Harris DN, Kessler MD, et al. Sequencing of 53,831 diverse genomes from the NHLBI TOPMed Program. Nature. 2021;590(7845):290–299. doi:10.1038/s41586-021-03205-y

23. Das S, Forer L, Schönherr S, Sidore C, Locke AE, Kwong A, et al. Next-generation genotype imputation service and methods. Nat Genet. 2016;48(10):1284–7

24. Chang CC, Chow CC, Tellier LC, Vattikuti S, Purcell SM, Lee JJ. Second-generation PLINK: rising to the challenge of larger and richer datasets. GigaScience. 2015;4(1). doi:10.1186/s13742-015-0047-8

25. Abraham G, Qiu Y, Inouye M. FlashPCA2: principal component analysis of Biobank-scale genotype datasets. Bioinformatics. 2017;33(17):2776–8.

26. Data | 1000 Genomes. Accessed January 31, 2023. https://www.internationalgenome.org/data

27. Auton A, Abecasis GR, Altshuler DM, et al. A global reference for human genetic variation. Nature. 2015;526(7571):68–74. doi:10.1038/nature15393

28. Samuel A. Lambert, Laurent Gil, Simon Jupp, Scott C. Ritchie, Yu Xu, Annalisa Buniello, Aoife McMahon, Gad Abraham, Michael Chapman, Helen Parkinson, John Danesh, Jacqueline A. L. MacArthur and Michael Inouye. The Polygenic Score Catalog as an open database for reproducibility and systematic evaluation Nature Geneticsdoi: 10.1038/s41588-021-00783-5 (2021).

29. Privé F, Arbel J, Vilhjálmsson BJ. LDpred2: better, faster, stronger [published online ahead of print, 2020 Dec 16]. Bioinformatics. 2020;36(22-23):5424–5431. doi:10.1093/bioinformatics/btaa1029

30. Denny JC, Bastarache L, Ritchie MD, et al. Systematic comparison of phenome-wide association study of electronic medical record data and genome-wide association study data. Nat Biotechnol. 2013;31(12):1102–1111. doi:10.1038/nbt.2749

31. The Python Language Reference. Python documentation. Accessed January 31, 2023. https://docs.python.org/3/reference/index.html

32. The Comprehensive R Archive Network. Accessed January 31, 2023. https://cran.r-project.org/

33. Services I of M (US) C on MA to PHC, Millman M. A Model for Monitoring Access. National Academies Press (US); 1993. Accessed January 31, 2023. https://www.ncbi.nlm.nih.gov/books/NBK235891/

34. Viechtbauer W. Conducting Meta-Analyses in R with the metafor Package. J Stat Softw. 2010;36:1–48. doi:10.18637/jss.v036.i03

35. Liu M, Jiang Y, Wedow R, et al. Association studies of up to 1.2 million individuals yield new insights into the genetic etiology of tobacco and alcohol use. Nat Genet. 2019;51(2):237–244. doi:10.1038/s41588-018-0307-5

36. Elsworth B, Lyon M, Alexander T, et al. The MRC IEU OpenGWAS data infrastructure. bioRxiv. Published online August 10, 2020. doi:10.1101/2020.08.10.244293

37. Hemani G, Zheng J, Elsworth B, et al. The MR-Base platform supports systematic causal inference across the human phenome. eLife. doi:10.7554/eLife.34408

38. Ding Y, Hou K, Burch KS, et al. Large uncertainty in individual polygenic risk score estimation impacts PRS-based risk stratification. Nat Genet. 2022;54(1):30–39. doi:10.1038/s41588-021-00961-5

39. Prevention (US) C for DC and, Promotion (US) NC for CDP and H, Health (US) O on S and. How Tobacco Smoke Causes Disease: The Biology and Behavioral Basis for Smoking-Attributable Disease. Centers for Disease Control and Prevention (US); 2010. Accessed January 31, 2023. https://www.ncbi.nlm.nih.gov/books/NBK53017/

40. Roy A, Rawal I, Jabbour S, et al. Tobacco and Cardiovascular Disease: A Summary of Evidence. In: Prabhakaran D, Anand S, Gaziano TA, et al., editors. Cardiovascular, Respiratory, and Related Disorders. 3rd edition. Washington (DC): The International Bank for Reconstruction and Development / The World Bank; 2017 Nov 17. Chapter 4. Available from: https://www.ncbi.nlm.nih.gov/books/NBK525170/ doi: 10.1596/978-1-4648-0518-9_ch4

41. De Angelis F, Wendt FR, Pathak GA, et al. Drinking and smoking polygenic risk is associated with childhood and early-adulthood psychiatric and behavioral traits independently of substance use and psychiatric genetic risk. Transl Psychiatry. 2021;11(1):1–12. doi:10.1038/s41398-021-01713-z

42. Carreras-Torres R, Johansson M, Haycock PC, et al. Role of obesity in smoking behaviour: Mendelian randomisation study in UK Biobank. BMJ. 2018;361. doi:10.1136/bmj.k1767

43. Thorgeirsson TE, Gudbjartsson DF, Sulem P, et al. A common biological basis of obesity and nicotine addiction. Transl Psychiatry. 2013;3(10):e308. doi:10.1038/tp.2013.81

