## Supplemental Figures and Tables for "Polygenic scores for tobacco use provide insights into systemic health risks in a diverse EHR-linked biobank in Los Angeles"

**Supplementary Table 1 - TUD-PGS association with TUD across GIAs**

| GIA | β | SE | Z | P>\|z\| | [0.025 | 0.975] | OR | OR_lower_CI | OR_upper_CI |
|---|---|---|---|---|---|---|---|---|---|
| European American | 0.18 | 0.02 | 10.44 | 1.66E-25 | 0.15 | 0.22 | 1.20 | 1.16 | 1.24 |
| Hispanic/Latin American | 0.17 | 0.04 | 4.73 | 2.24E-06 | 0.10 | 0.24 | 1.19 | 1.11 | 1.28 |
| East Asian American | 0.17 | 0.05 | 3.10 | 1.93E-03 | 0.06 | 0.27 | 1.18 | 1.06 | 1.30 |
| African American | 0.04 | 0.06 | 0.66 | 5.07E-01 | -0.08 | 0.16 | 1.04 | 0.93 | 1.17 |

**Supplementary Table 2 - TUD-PGS association with TUD across quintiles and GIAs**

| PGS_ Quantile | Coeff | SE | Z | P>\|z\| | [0.025 | 0.975] | OR | OR_ lower_CI | OR_ upper_CI | GIA |
|---|---|---|---|---|---|---|---|---|---|---|
| 2 | 0.10 | 0.06 | 1.86 | 6.27E-02 | -0.005 | 0.21 | 1.11 | 0.99 | 1.24 | European American |
| 3 | 0.22 | 0.06 | 3.95 | 7.70E-05 | 0.11 | 0.33 | 1.25 | 1.12 | 1.39 | European American |
| 4 | 0.31 | 0.06 | 5.58 | 2.47E-08 | 0.20 | 0.42 | 1.36 | 1.22 | 1.52 | European American |
| 5 | 0.52 | 0.06 | 9.45 | 3.25E-21 | 0.41 | 0.63 | 1.69 | 1.51 | 1.88 | European American |
| 2 | 0.15 | 0.12 | 1.32 | 1.86E-01 | -0.07 | 0.38 | 1.17 | 0.93 | 1.47 | Hispanic/ Latin American |
| 3 | 0.16 | 0.12 | 1.39 | 1.66E-01 | -0.07 | 0.39 | 1.18 | 0.94 | 1.48 | Hispanic/ Latin American |
| 4 | 0.29 | 0.12 | 2.53 | 1.13E-02 | 0.07 | 0.52 | 1.34 | 1.07 | 1.68 | Hispanic/ Latin American |
| 5 | 0.54 | 0.12 | 4.65 | 3.27E-06 | 0.31 | 0.76 | 1.71 | 1.36 | 2.14 | Hispanic/ Latin American |
| 2 | 0.19 | 0.17 | 1.12 | 2.64E-01 | -0.15 | 0.53 | 1.21 | 0.86 | 1.70 | East Asian American |
| 3 | 0.52 | 0.17 | 3.1 | 1.94E-03 | 0.19 | 0.85 | 1.69 | 1.21 | 2.35 | East Asian American |

| | | | | | | | | | | |
|---|---|---|---|---|---|---|---|---|---|---|
| 4 | 0.31 | 0.17 | 1.82 | 6.87E-02 | -0.02 | 0.65 | 1.37 | 0.98 | 1.91 | East Asian American |
| 5 | 0.47 | 0.17 | 2.76 | 5.77E-03 | 0.14 | 0.80 | 1.60 | 1.15 | 2.23 | East Asian American |
| 2 | -0.10 | 0.18 | -0.59 | 5.58E-01 | -0.45 | 0.24 | 0.90 | 0.63 | 1.27 | African American |
| 3 | 0.01 | 0.18 | 0.06 | 9.49E-01 | -0.34 | 0.36 | 1.01 | 0.71 | 1.43 | African American |
| 4 | 0.12 | 0.18 | 0.69 | 4.93E-01 | -0.23 | 0.48 | 1.13 | 0.79 | 1.62 | African American |
| 5 | 0.02 | 0.18 | 0.10 | 9.18E-01 | -0.34 | 0.38 | 1.02 | 0.71 | 1.47 | African American |

**Supplemental Figure 1a**
**DAG showing the relationship evaluated in the PGS-PheWAS meta-analysis**

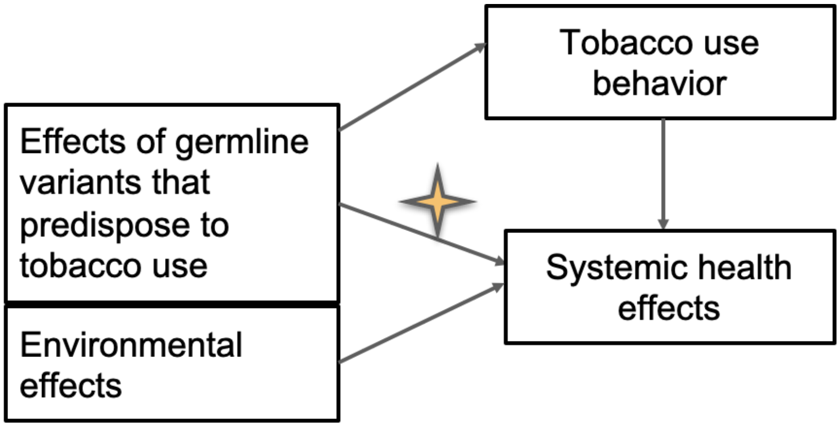


**Supplementary Table 3 - Significant associations between TUD-PGS and 1847 traits in the PGS-PheWAS cross-ancestry meta-analysis**

| Phecode | beta | SE | Z | P Value | CI.LB | CI.UB | QEp | Phenotype | Category |
|---|---|---|---|---|---|---|---|---|---|
| 278.11 | 0.12 | 0.02 | 6.06 | 1.38E-09 | 0.08 | 0.17 | 0.46 | Morbid obesity | endocrine/metabolic |
| 496.21 | 0.25 | 0.04 | 5.95 | 2.73E-09 | 0.17 | 0.33 | 0.10 | Obstructive chronic bronchitis | respiratory |
| 316 | 0.12 | 0.02 | 5.47 | 4.45E-08 | 0.08 | 0.16 | 0.56 | Substance addiction and disorders | mental disorders |

| | | | | | | | | | |
|---|---|---|---|---|---|---|---|---|---|
| 411 | 0.09 | 0.02 | 5.24 | 1.61E-07 | 0.05 | 0.12 | 0.81 | Ischemic Heart Disease | circulatory system |
| 228.1 | -0.10 | 0.02 | -5.09 | 3.49E-07 | -0.14 | -0.06 | 0.74 | Hemangioma of skin and subcutaneous tissue | neoplasms |
| 428.1 | 0.12 | 0.02 | 5.03 | 4.80E-07 | 0.07 | 0.16 | 0.33 | Congestive heart failure (CHF) NOS | circulatory system |
| 327.3 | 0.08 | 0.02 | 5.02 | 5.29E-07 | 0.05 | 0.11 | 0.23 | Sleep apnea | neurological |
| 228 | -0.09 | 0.02 | -4.78 | 1.74E-06 | -0.13 | -0.05 | 0.76 | Hemangioma and lymphangioma, any site | neoplasms |
| 411.3 | 0.12 | 0.03 | 4.53 | 5.83E-06 | 0.07 | 0.17 | 0.71 | Angina pectoris | circulatory system |
| 530 | 0.06 | 0.01 | 4.46 | 8.05E-06 | 0.03 | 0.09 | 0.31 | Diseases of esophagus | digestive |
| 428 | 0.09 | 0.02 | 4.42 | 9.57E-06 | 0.05 | 0.14 | 0.99 | Congestive heart failure; nonhypertensive | circulatory system |
| 338.2 | 0.06 | 0.01 | 4.41 | 1.05E-05 | 0.03 | 0.09 | 0.38 | Chronic pain | neurological |
| 509 | 0.09 | 0.02 | 4.37 | 1.24E-05 | 0.04817 | 0.13 | 0.96 | Respiratory failure, insufficiency, arrest | respiratory |
| 278.1 | 0.11 | 0.03 | 4.34 | 1.42E-05 | 0.064552 | 0.17 | 0.12 | Obesity | endocrine/metabolic |
| 411.2 | 0.11 | 0.03 | 4.34 | 1.45E-05 | 0.060558 | 0.16 | 0.67 | Myocardial infarction | circulatory system |
| 300 | 0.06 | 0.01 | 4.27 | 1.95E-05 | 0.031596 | 0.09 | 0.87 | Anxiety disorders | mental disorders |
| 508 | 0.07 | 0.02 | 4.24 | 2.23E-05 | 0.035846 | 0.097 | 0.53 | Pulmonary collapse; interstitial and compensatory emphysema | respiratory |

**Supplemental Figure 1b**
**DAG showing the relationship evaluated in the PGS-PheWAS never-smoker analysis**

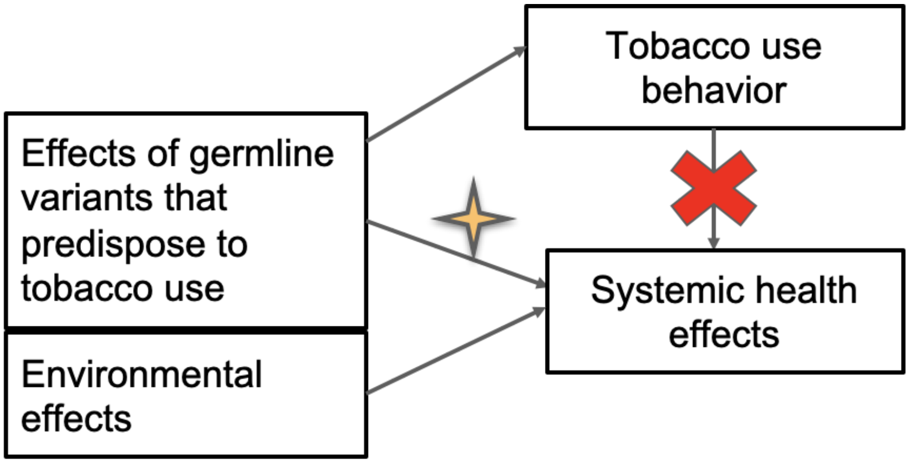


**Supplementary Table 4 - Significant associations between TUD-PGS and 1847 traits in the 'never-smoker' PGS-PheWAS in EA ancestry group**

| Phecode | Coef | SE | Z | P Value | [0.025 | 0.975] | Phenotype | Category |
|---|---|---|---|---|---|---|---|---|
| 278.1 | 0.13 | 0.03 | 5.09 | 3.54E-07 | 0.08 | 0.18 | Obesity | endocrine/metabolic |
| 317 | 0.23 | 0.05 | 4.8 | 1.61E-06 | 0.14 | 0.32 | Alcohol-related disorders | mental disorders |
| 721 | 0.12 | 0.03 | 4.79 | 1.64E-06 | 0.074 | 0.17 | Spondylosis and allied disorders | musculoskeletal |
| 278.11 | 0.16 | 0.03 | 4.70 | 2.56E-06 | 0.09 | 0.23 | Morbid obesity | endocrine/metabolic |
| 150 | 0.71 | 0.15 | 4.67 | 3.05E-06 | 0.41 | 1.01 | Cancer of esophagus | neoplasms |
| 228 | -0.12 | 0.03 | -4.58 | 4.67E-06 | -0.17 | -0.07 | Hemangioma and lymphangioma, any site | neoplasms |
| 317.1 | 0.23 | 0.05 | 4.29 | 1.78E-05 | 0.12 | 0.33 | Alcoholism | mental disorders |
| 401 | 0.09 | 0.02 | 4.20 | 2.62E-05 | 0.05 | 0.14 | Hypertension | circulatory system |

**Supplemental Figure 2**
**TUD-PGS association with Lung Cancer across PGS quintiles among ever smokers and never smokers**
Associations between TUD-PGS and lung cancer. X-axis represents the top 4 quintiles grouped by TUD-PGS. Y axis represents effect sizes represented by odds ratios. Red line indicates OR =1. A reverse trend from alcohol-related disorders and obesity is noted in lung cancer demonstrating a trend of higher ORs in ever-smokers compared to never-smokers, across the quintiles going from lowest to highest.

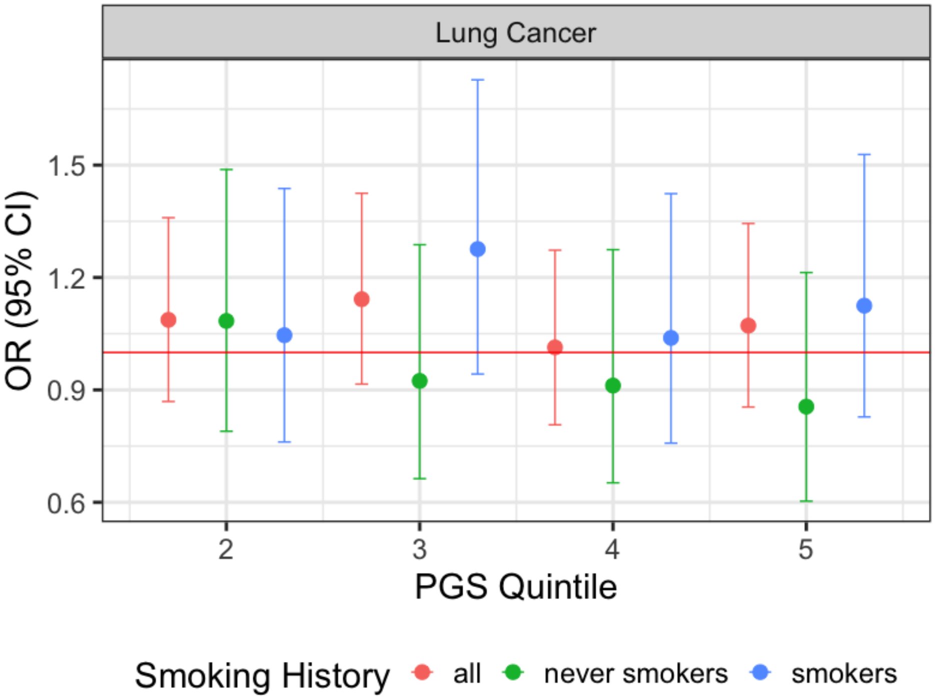


**Supplementary Table 5 - Associations between Alcohol-Related Disorders, Obesity, and Lung cancer and PGS quantiles traits in the 'ever-smoker' and 'never-smoker' groups**

| PGS_ Quantile | Coef. | SE | Z | P>\|z\| | [0.025 | 0.975] | Phecode | Smoking History | OR | OR_ Lower_ CI | OR_ Upper_ CI | Phenotype |
|---|---|---|---|---|---|---|---|---|---|---|---|---|
| 2 | 0.004 | 0.05 | 0.08 | 9.40E-01 | -0.095 | 0.10 | 278.1 | all | 1.00 | 0.91 | 1.11 | Obesity |
| 3 | 0.16 | 0.05 | 3.31 | 9.38E-04 | 0.07 | 0.26 | 278.1 | all | 1.18 | 1.07 | 1.30 | Obesity |
| 4 | 0.19 | 0.05 | 3.87 | 1.11E-04 | 0.09 | 0.29 | 278.1 | all | 1.21 | 1.1 | 1.33 | Obesity |
| 5 | 0.28 | 0.05 | 5.61 | 2.02E-08 | 0.18 | 0.37 | 278.1 | all | 1.32 | 1.2 | 1.45 | Obesity |
| 2 | 0.04 | 0.09 | 0.50 | 6.17E-01 | -0.13 | 0.22 | 278.1 | smokers | 1.05 | 0.88 | 1.25 | Obesity |
| 3 | 0.11 | 0.09 | 1.29 | 1.97E-01 | -0.06 | 0.28 | 278.1 | smokers | 1.12 | 0.94 | 1.33 | Obesity |
| 4 | 0.23 | 0.09 | 2.73 | 6.34E-03 | 0.07 | 0.40 | 278.1 | smokers | 1.26 | 1.07 | 1.49 | Obesity |
| 5 | 0.16 | 0.08 | 1.87 | 6.22E-02 | -0.008 | 0.32 | 278.1 | smokers | 1.17 | 0.99 | 1.38 | Obesity |
| 2 | -0.02 | 0.06 | -0.40 | 6.93E-01 | -0.15 | 0.1 | 278.1 | never smokers | 0.98 | 0.86 | 1.10 | Obesity |
| 3 | 0.18 | 0.06 | 2.95 | 3.22E-03 | 0.06 | 0.30 | 278.1 | never smokers | 1.20 | 1.06 | 1.35 | Obesity |
| 4 | 0.15 | 0.06 | 2.4 | 1.67E-02 | 0.027 | 0.27 | 278.1 | never smokers | 1.16 | 1.03 | 1.31 | Obesity |

| | | | | | | | | | | | | |
|---|---|---|---|---|---|---|---|---|---|---|---|---|
| 5 | 0.32 | 0.06 | 5.33 | 9.73E-08 | 0.20 | 0.44 | 278.1 | never smokers | 1.38 | 1.23 | 1.56 | Obesity |
| 2 | 0.06 | 0.08 | 0.72 | 4.69E-01 | -0.10 | 0.22 | 317 | all | 1.06 | 0.90 | 1.25 | Alcohol Related Disorders |
| 3 | 0.11 | 0.08 | 1.39 | 1.63E-01 | -0.05 | 0.27 | 317 | all | 1.12 | 0.95 | 1.32 | Alcohol Related Disorders |
| 4 | 0.15 | 0.08 | 1.84 | 6.60E-02 | -0.009 | 0.31 | 317 | all | 1.16 | 0.99 | 1.36 | Alcohol Related Disorders |
| 5 | 0.33 | 0.08 | 4.22 | 2.44E-05 | 0.18 | 0.49 | 317 | all | 1.39 | 1.19 | 1.63 | Alcohol Related Disorders |
| 2 | -0.05 | 0.12 | -0.47 | 6.41E-01 | -0.29 | 0.18 | 317 | smokers | 0.95 | 0.75 | 1.19 | Alcohol Related Disorders |
| 3 | 0.07 | 0.11 | 0.66 | 5.09E-01 | -0.15 | 0.30 | 317 | smokers | 1.08 | 0.86 | 1.35 | Alcohol Related Disorders |
| 4 | -0.04 | 0.11 | -0.38 | 7.08E-01 | -0.27 | 0.18 | 317 | smokers | 0.96 | 0.77 | 1.20 | Alcohol Related Disorders |
| 5 | 0.11 | 0.11 | 0.99 | 3.24E-01 | -0.11 | 0.32 | 317 | smokers | 1.11 | 0.90 | 1.38 | Alcohol Related Disorders |
| 2 | 0.11 | 0.12 | 0.95 | 3.42E-01 | -0.12 | 0.35 | 317 | never smokers | 1.12 | 0.88 | 1.41 | Alcohol Related Disorders |
| 3 | 0.03 | 0.12 | 0.27 | 7.85E-01 | -0.21 | 0.27 | 317 | never smokers | 1.03 | 0.81 | 1.31 | Alcohol Related Disorders |
| 4 | 0.22 | 0.12 | 1.85 | 6.42E-02 | -0.01 | 0.45 | 317 | never smokers | 1.24 | 0.99 | 1.56 | Alcohol Related Disorders |
| 5 | 0.39 | 0.11 | 3.37 | 7.64E-04 | 0.16 | 0.61 | 317 | never smokers | 1.47 | 1.17 | 1.84 | Alcohol Related Disorders |
| 2 | 0.08 | 0.11 | 0.73 | 4.65E-01 | -0.14 | 0.31 | 165.1 | all | 1.09 | 0.87 | 1.36 | Lung Cancer |
| 3 | 0.13 | 0.11 | 1.18 | 2.39E-01 | -0.09 | 0.35 | 165.1 | all | 1.14 | 0.91 | 1.42 | Lung Cancer |
| 4 | 0.01 | 0.11 | 0.11 | 9.08E-01 | -0.21 | 0.24 | 165.1 | all | 1.01 | 0.81 | 1.27 | Lung Cancer |

| | | | | | | | | | | | | |
|---|---|---|---|---|---|---|---|---|---|---|---|---|
| 5 | 0.07 | 0.11 | 0.60 | 5.50E-01 | -0.16 | 0.30 | 165.1 | all | 1.07 | 0.85 | 1.34 | Lung Cancer |
| 2 | 0.05 | 0.16 | 0.28 | 7.82E-01 | -0.27 | 0.36 | 165.1 | smokers | 1.05 | 0.76 | 1.44 | Lung Cancer |
| 3 | 0.24 | 0.15 | 1.58 | 1.15E-01 | -0.06 | 0.55 | 165.1 | smokers | 1.28 | 0.94 | 1.73 | Lung Cancer |
| 4 | 0.04 | 0.16 | 0.24 | 8.13E-01 | -0.28 | 0.35 | 165.1 | smokers | 1.04 | 0.76 | 1.42 | Lung Cancer |
| 5 | 0.11 | 0.16 | 0.75 | 4.52E-01 | -0.19 | 0.42 | 165.1 | smokers | 1.12 | 0.83 | 1.53 | Lung Cancer |
| 2 | 0.08 | 0.16 | 0.50 | 6.18E-01 | -0.24 | 0.40 | 165.1 | never smokers | 1.08 | 0.79 | 1.49 | Lung Cancer |
| 3 | -0.08 | 0.17 | -0.47 | 6.40E-01 | -0.41 | 0.25 | 165.1 | never smokers | 0.92 | 0.66 | 1.29 | Lung Cancer |
| 4 | -0.09 | 0.17 | -0.54 | 5.88E-01 | -0.43 | 0.24 | 165.1 | never smokers | 0.91 | 0.65 | 1.27 | Lung Cancer |
| 5 | -0.16 | 0.18 | -0.88 | 3.81E-01 | -0.51 | 0.19 | 165.1 | never smokers | 0.86 | 0.60 | 1.21 | Lung Cancer |

**Supplemental Figure 3**

**MR results between cigarettes smoked per day (GSCAN Consortium) and waist circumference (MRC-UBristol and UKBB) and body mass index (UKBB)**

Mendelian Randomization results across multiple MR methods using summary statistics for cigarettes smoked per day from the GSCAN Consortium and for waist circumference from MRC-UBristol and body mass index from UKBB. X axis represents the effect sizes and Y axis represents the MR method used.

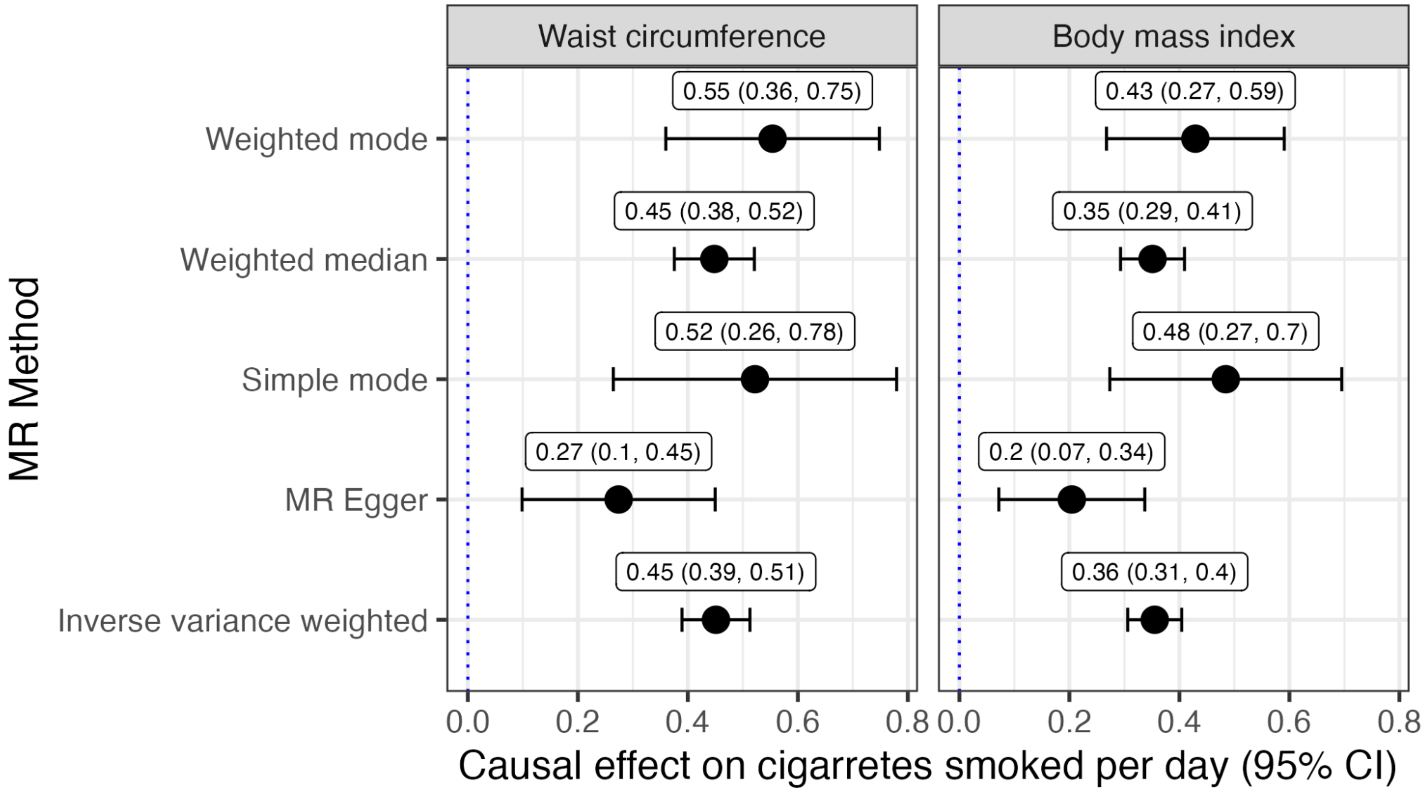
